# Impact of Midwifery-led units in Spain: lessons from the first 5 years

**DOI:** 10.1101/2024.07.16.24310479

**Authors:** Roser Palau-Costafreda, Lluna Orus-Covisa, Edgar Vicente-Castellví, Xavier Espada-Trespalacios, Albert Medina Català, Carlota Alcover, Noemí Obregón Gutiérrez, Ramon Escuriet

## Abstract

**Introduction:** The global rise in medical interventions during childbirth, such as caesarean sections, has raised concerns regarding their necessity and impact on maternal and neonatal outcomes. Midwifery-led units (MLUs) have demonstrated lower intervention rates and higher maternal satisfaction.This study evaluates the implementation and effects of the first MLU in the Spanish National Health System.

**Methods:** A retrospective cross-sectional trend study and a cohort study were conducted to compare childbirth interventions and outcomes at XX with other hospitals of varying complexities.

**Results:** The introduction of the MLU at XX resulted in a significant reduction in caesarean sections, decreasing from 23.5% to 13.5%, and an increase in spontaneous vaginal births, rising from 64.2% to 78.7%. These trends reversed following the MLU’s closure in 2022, with caesarean sections increasing to 22.9% and spontaneous births dropping to 69.0%. The MLU served 1286 women, with the majority classified as low-risk pregnancies. Obstetric emergencies in the MLU were low and comparable to those in countries with established MLUs.

**Discussion:** This study highlights the potential benefits of integrating MLUs into traditionally medicalized healthcare systems to promote physiological childbirth and reduce unnecessary interventions. The positive outcomes achieved at HM are comparable to those in countries with more established MLU practices, reflecting the unit’s commitment to evidence-based care. The increasing interest among women in midwifery-led care indicates a broader demand for supportive, less medicalized childbirth environments.

**Conclusions:** MLU can lead to lower caesarean section rates and higher spontaneous vaginal birth rates, contributing to more positive maternal and neonatal outcomes. However, sustained support and investment in these units are crucial to maintain these benefits. Policymakers and healthcare providers should consider expanding the integration of MLUs within the Spanish National Health System to enhance maternal care quality and align with best practices.

## 1. INTRODUCTION

The global rise in medical interventions during childbirth, such as caesarean sections and medical inductions, is a significant concern for health systems worldwide (Boerma et al., 2018; Dumont & Guilmoto, 2020; Opiyo et al., 2019; Vogel et al., 2015; WHO, 2018). Improvements in maternal or neonatal mortality rates have not consistently followed the increase in childbirth intervention rates (WHO, 2023). Rather, the routine use of these interventions not only increases the consumption of healthcare resources but also poses potential health risks for both women and newborns (Sandall et al., 2018), suggesting a trend of “too much, too soon” interventions that may not be medically necessary (Miller et al., 2016).

In this context, traditional Obstetric Units (OUs), which have long dominated the field of childbirth, are under the spotlight for their highly medicalised approach (Hodnett et al., 2012; Renfrew et al., 2014). Despite their aim to reduce maternal and neonatal mortality through technological and procedural interventions, evidence suggests that OUs may not provide optimal conditions for low-risk pregnancies (Fikre et al., 2023). Alternatively, Midwifery-Led Units (MLUs) emerge as a viable option. Unlike OUs, MLUs emphasise minimal intervention and support physiological childbirth processes in home-like settings. Evidence from countries such as the United Kingdom, Australia, and Canada suggests that MLUs achieve lower intervention rates (Hodnett et al., 2012; Scarf et al., 2018), higher maternal satisfaction (Hodnett et al., 2012), and more favourable economic outcomes due to reduced clinical procedures (Bernitz et al., 2012) and shorter hospital stays (Sandall et al., 2018). Accordingly, health systems are increasingly supporting these units, albeit at different paces.

In Spain, a country with a traditionally obstetrician-led approach to maternity care and limited integration of midwifery services, the first MLU opened in 2017 in Catalonia, one of the country’s 17 regions with autonomy to adapt healthcare delivery, including maternity services, to its unique setting (Bernal-Delgado et al., 2018). In this region, antenatal and postnatal services are universally accessible at Sexual and Reproductive Healthcare Centres (ASSIRs), which are connected to primary care. Childbirth and immediate postpartum care are delivered in maternity hospitals, categorised into levels (I-III) according to their capacity to provide varying complexities of maternal and neonatal care (Figure 1). Women with uncomplicated pregnancies receive autonomous care from midwifery professionals as part of a multidisciplinary team at level-I and -II hospitals, while very-high-complexity pregnancies receive specialised medical attention within a collaborative team framework at level-III hospitals (Generalitat de Catalunya, 2020).

**Fig. 1.**
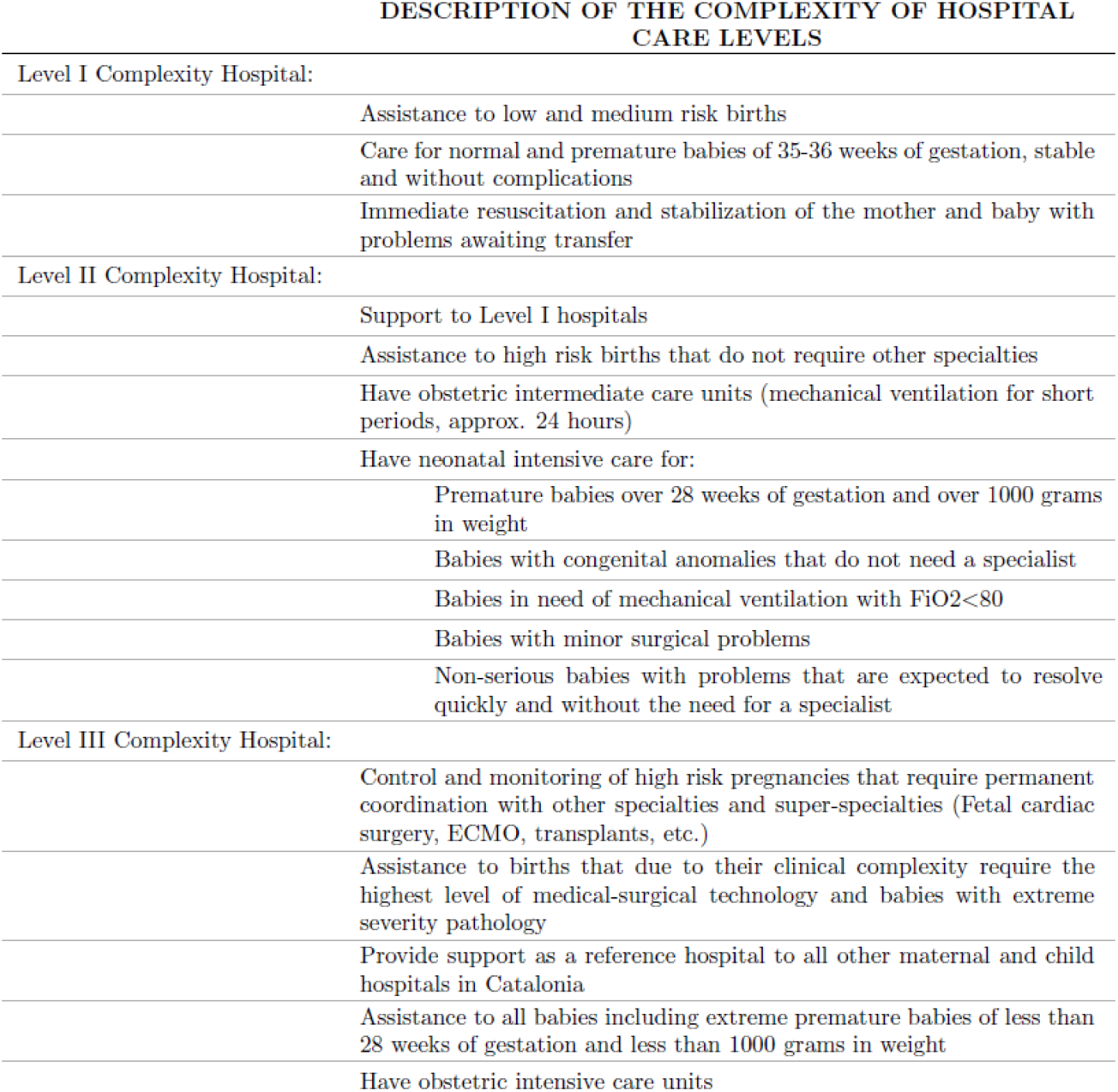
Description of the complexity of hospital care levels

When the MLU was implemented, in 2017, Catalonia accounted for almost 17% of Spain’s births, 27.8% of which were by caesarean section (Idescat, 2018). This was slightly above the national average (26.5%) (INE, 2018) and well above Europe’s leading countries such as Finland (16.5%) or Sweden (16.6%) (Eurostat, 2022). However, there were substantial variations across centres, with the share of caesarean births ranging from 10.4% to 62.8% in level-I complexity hospitals (Observatori del Sistema de Salut de Catalunya, 2018).

Against this background, the Catalan Department of Health, as part of a broader strategy to promote positive births (Generalitat de Catalunya, 2021), introduced the first MLU within Hospital XX (HM), a level-I complexity hospital with one of the highest proportions of caesarean sections in Catalonia (Observatori del Sistema de Salut de Catalunya, 2018). HM is a community public-private centre located 30 km away from Barcelona that serves a population of about 160,000 people (FHSJDM, 2024a) and manages an average of 650 childbirths per year (FHSJDM, 2024b). As a level I-complexity hospital, it attends women with uncomplicated pregnancies between 35–42 weeks of gestation. Ever since the implementation of the MLU, women interested in midwifery-led care were scheduled for an appointment with the MLU team, during which their pregnancy risk was assessed according to national and international guidelines (NICE, 2014). Based on this assessment, they were either admitted to the MLU or transferred to HM’s or other hospitals’ OUs. Women admitted to the MLU were provided continuous care from the small team throughout the end of pregnancy, labour and postpartum unless complications or risk factors arose, logistic issues emerged (e.g. MLU not available for birth) or the woman requested epidural analgesia during labour. They were then discharged after 24 hours of delivery unless maternal or neonatal complications arose.

During the COVID-19 pandemic, both the MLU and HM’s OU remained open, unlike other OUs in the region, which were temporarily converted to COVID-19-dedicated units. Nevertheless, the MLU closed at the end of 2022 due to organisational factors. Although it reopened at the beginning of 2024, the closure of the MLU presents a unique case study for evaluating the broader implications of introducing such a unit for maternity care in Spain, as well as contributing to the ongoing discussion on reducing childbirth interventions and enhancing the quality of care.

Therefore, this study aims to evaluate the implementation of Spain’s first alongside MLU over its initial five years of operation, from 2017 to 2022. By examining maternal and neonatal outcomes alongside childbirth intervention trends, this study seeks to provide valuable insights into the benefits and challenges of integrating MLUs into a traditionally medicalized healthcare context.

## 2. MATERIALS AND METHODS

We conducted a retrospective cross-sectional trend study of childbirths attended in Catalonia, Spain, between 2018 and 2023, combined with a retrospective cohort study of all women who opted for midwifery-led care at the study site, HM, from December 2017 to December 2022.

In a first phase of the study, we aimed to compare HM’s childbirth intervention trends with those of level I-, II- and III-complexity hospitals from 2018, right after the MLU was first opened, to 2023, a year after the closure of the MLU. For this purpose, we drew data from the Minimum Basic Data Set (MBDS) of the Catalan Health Service, an administrative register subject to Ministerial regulation that compiles population-based data on healthcare utilisation from all publicly financed health institutions in Catalonia (Generalitat de Catalunya, 2024). The MBDS records all interactions of citizens with public healthcare services, combining their sociodemographic and clinical information with variables related to the centres where they were attended, as notified by the latter. Clinical information is classified according to the International Classification of Diseases 10th Revision Clinical Modification (ICD-10-CM), which allows for the codification of pathological and non-pathological situations, including pregnancies. Hence, we retrieved childbirth information from HM as well as from other health institutions aggregated by level of complexity. Data were analysed by means of descriptive statistics and summarised in figures for visual examination.

In a second phase, we delved further into these trends by assessing the interest in, utilisation of, and outcomes delivered by the MLU’s first five years of functioning. We retrieved individual-level data of all women who contacted the MLU from December 2017 to December 2022, regardless of whether they were finally admitted to the MLU. This made up a total of 1,286 women. Data were collected by the MLU team as women contacted the unit and complemented with information from the hospital’s system. The final dataset includes women’s socio-demographic characteristics; pre-existing medical conditions; obstetric history; gynaecological characteristics; type of birth; maternal outcomes; neonatal outcomes; and transfers to other units. Studied variables were residential area (within or outside the hospital’s coverage area), initial pregnancy risk assessment (low, medium, high), pathway of care (no transfer, antenatal transfer, intrapartum transfer, postpartum transfer), type of birth (caesarean section, instrumental birth, spontaneous vaginal birth with epidural, spontaneous vaginal birth outside the water, spontaneous vaginal birth with water immersion), emergency during labour at the MLU (yes, no), type of emergency (neonatal resuscitation, shoulder dystocia, postpartum haemorrhage), transfer to a neonatal intensive care unit (yes, no), and 5-minute Apgar score (<7, ≥7). Year of birth and parity (nulliparous, multiparous) were used as stratification variables. These were described using descriptive statistics according to the nature of each variable. Statistics were performed using R v4.2.2 (R Core Team, 2022).

Ethical approval was granted by the Fundació Unió Catalana d’Hospitals Ethics Committee (Ref CEI 21/03).

## 3. RESULTS

Figure 2 shows the annual distribution of childbirths in Catalonia, classified by type of birth and hospital level (see Supplementary Tables 1 and 2 for further details), from 2018 to 2023. During this period, spontaneous births were the most frequent in all hospital levels, with proportions ranging from 64% to 70% approximately. Level-I hospitals had the highest shares of spontaneous births, followed by level-II and level-III hospitals, respectively. Instrumental birth percentages ranged from 10% to 12% across all hospital levels, showing a modest decrease throughout the studied period. The proportion of caesarean sections ranged from 19% to 23% in level-I and -II hospitals, and from 25% to 27% in level-III hospitals. Unlike instrumental births, caesarean sections slightly increased in all hospital levels between 2018 and 2023.

**Fig. 2.**
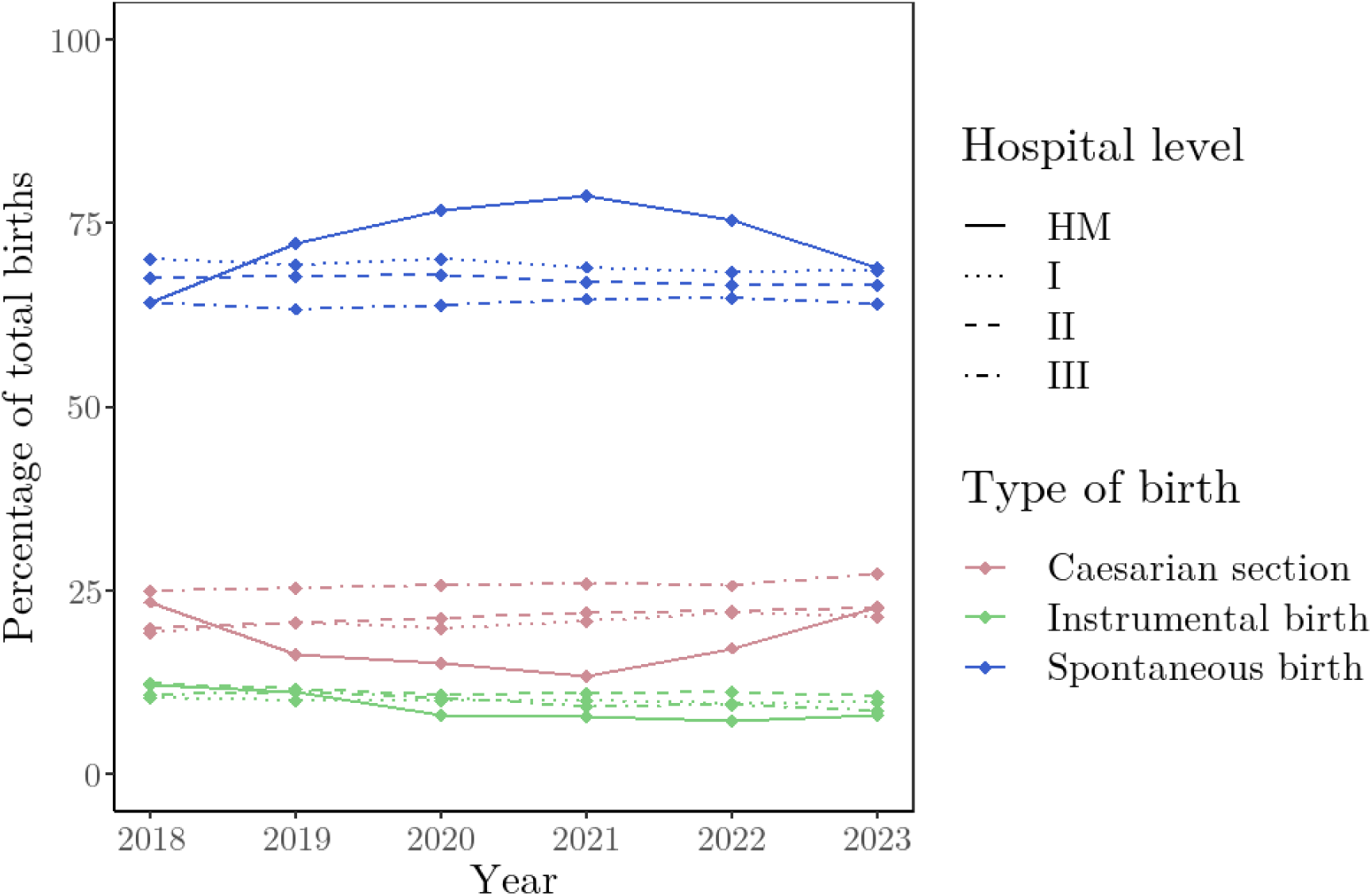
Type of birth trends across hospital levels and Hospital XX (HM). 2018-2023. Source: own elaboration based on data from the Minimum Basic Data Set of the Catalan Health Service.

In contrast, HM shows a different pattern. At the beginning of the study period, right after the implementation of the MLU, HM’s share of spontaneous births (64.2%) resembled that of level-III (64.3%) rather than level-I hospitals (70.1%), and the same occurred with the proportion of caesarean sections (23.5%, 24.9%, and 19.4%, respectively). Yet, from 2019 to 2021, the trends reversed, with caesarean sections falling to 13.5% and spontaneous births reaching 78.7%. This positive trend reversed again from 2022 to 2023, coinciding with the closure of the MLU: while caesarean sections rose back to 22.9%, spontaneous births decreased to 69.0%, meeting the proportions of other level-I hospitals (21.5% and 68.6%, respectively).

Focusing specifically on the implementation of the MLU, a total of 1,286 women contacted the unit for a first visit during the study period, showing an upward trend from 2018^1^ to 2022 (Table 1 and Figure 3): the number of women contacting the MLU grew from 110 in 2018 to 377 in 2022 and peaked at 409 in 2021, which strongly contrasts with the 650 childbirths on average attended at the hospital per year (FHSJDM, 2024b). Of these 1,286 women, 49 opted for the MLU in more than one pregnancy, suggesting satisfaction and trust in the service, and 958 (74.5%) were from outside the hospital’s coverage area, indicating an unmet need for more supportive approaches to childbirth (Table 1).

**Fig. 3.**
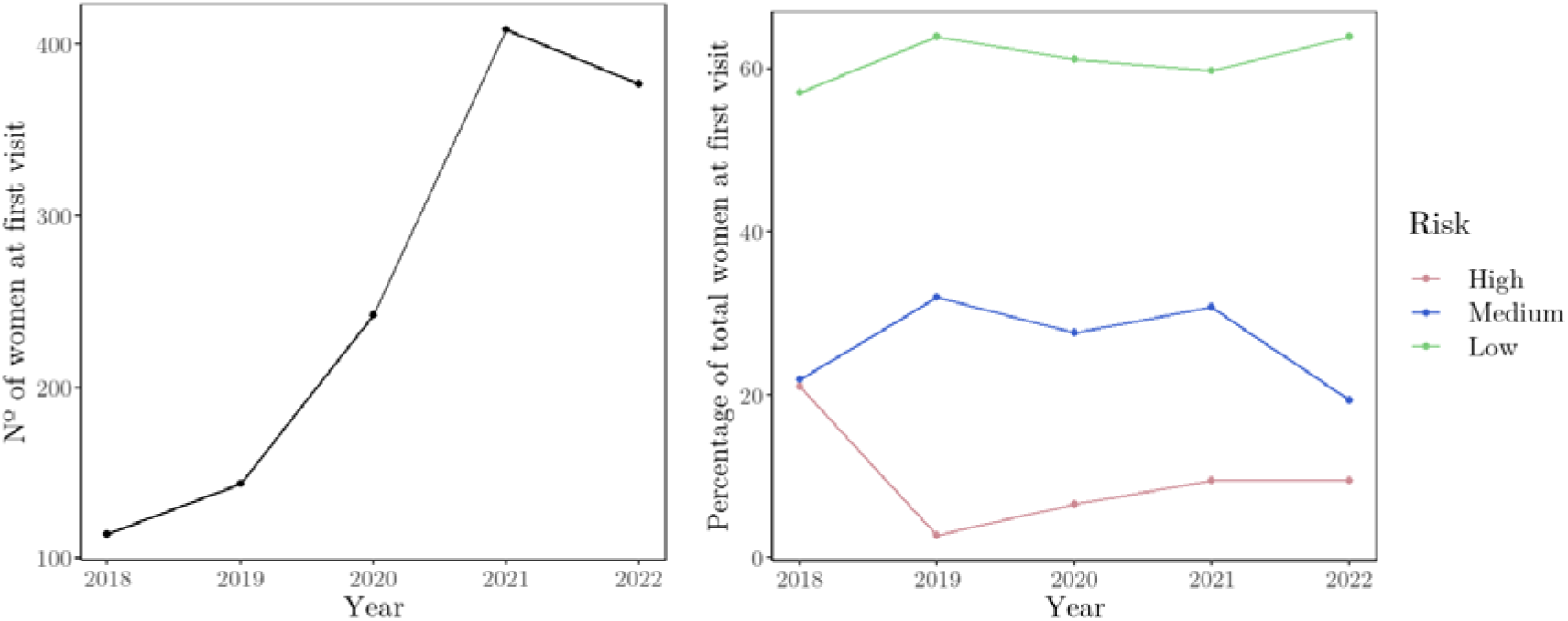
Trends of women attending a first visit at the Midwifery-Led Unit (left) and initial pregnancy risk assessment (right) from 2018 to 2022.

**Table 1.** Demographic and clinical characteristics by year (2017-2022)

|  | Total | 2017-2018 | 2019 | 2020 | 2021 | 2022 |
| --- | --- | --- | --- | --- | --- | --- |
|  | N=1286 | N=114 | N=144 | N=242 | N=409 | N=377 |
| Parity: |  |  |  |  |  |  |
| Nulliparous | 833 (64.8%) | 63 (55.3%) | 84 (58.3%) | 155 (64.0%) | 291 (71.1%) | 240 (63.7%) |
| Multiparous | 416 (32.3%) | 51 (44.7%) | 60 (41.7%) | 74 (30.6%) | 118 (28.9%) | 113 (30.0%) |
| Age: |  |  |  |  |  |  |
| <=29 | 242 (18.8%) | 22 (19.3%) | 32 (22.2%) | 48 (19.8%) | 65 (15.9%) | 75 (19.9%) |
| 30-34 | 506 (39.3%) | 47 (41.2%) | 56 (38.9%) | 82 (33.9%) | 171 (41.8%) | 150 (39.8%) |
| 35-39 | 390 (30.3%) | 41 (36.0%) | 46 (31.9%) | 77 (31.8%) | 117 (28.6%) | 109 (28.9%) |
| >=40 | 95 (7.39%) | 4 (3.51%) | 8 (5.56%) | 18 (7.44%) | 39 (9.54%) | 26 (6.90%) |
| Zone: |  |  |  |  |  |  |
| No | 958 (74.5%) | 79 (69.3%) | 77 (53.5%) | 167 (69.0%) | 345 (84.4%) | 290 (76.9%) |
| Yes | 313 (24.3%) | 35 (30.7%) | 66 (45.8%) | 66 (27.3%) | 64 (15.6%) | 82 (21.8%) |
| Risk in first visit: |  |  |  |  |  |  |
| High | 119 (9.25%) | 24 (21.1%) | 4 (2.78%) | 16 (6.61%) | 39 (9.54%) | 36 (9.55%) |
| Medium | 337 (26.2%) | 25 (21.9%) | 46 (31.9%) | 67 (27.7%) | 126 (30.8%) | 73 (19.4%) |
| Low | 790 (61.4%) | 65 (57.0%) | 92 (63.9%) | 148 (61.2%) | 244 (59.7%) | 241 (63.9%) |

As regards the socio-demographic characteristics of these women (Table 1), the most common age range (39.3%) was 30-34 years. The group aged between 35 and 39 years old was also substantial (30.3%). Those younger than 30 years and older than 39 years constituted 18.8% and 7.39%, respectively, and women aged 40+ years were the least frequent, likely due to increased labour complications associated with higher maternal ages. Regarding parity, nulliparous women were the most frequent, making up 64.8% of the 1,286 women attending a first visit.

Figure 3 shows the trends in initial pregnancy risk assessment. The majority of women that were scheduled for a first visit with the MLU team were classified as low risk, with percentages ranging from 57.0% in 2018 to 63.9% in 2019 and 2022 (Table 1). The percentage of medium-risk pregnancies fluctuated over the years, with higher values reached in 2019 (31.9%) and 2021 (30.8%), right amidst the MLU’s functioning period, and lower values shown during the opening and closing periods, in 2018 (21.9%) and 2022 (19.4%). Finally, the percentage of high-risk pregnancies was 21.1% in 2018. In 2019, it decreased drastically to 2.8%, but gradually increased again in the following years, reaching 9.6% in 2022.

Women followed different pathways of care (Figure 4). Of the 1,286 women who booked a first visit, 180 were excluded either during the first visit or throughout antenatal care, representing 14.0% of the initial cohort. Additionally, 38.5% of women were excluded before active labour, resulting in almost half of the women who initially planned to give birth at the MLU actually accessing it. Among those who did not initiate labour at the MLU, 22.1% were excluded due to distance reasons, indicating that geographical accessibility was a major barrier, 19.2% due to spontaneous rupture of membranes >24 hours, 17.9% because they requested epidural analgesia before active labour, 14.6% required medical induction, 11.3% faced organisational barriers such as staff shortages, 7.9% were suspected of foetal well-being loss, 6.5% had too rapidly progressing labours, and 0.6% presented maternal complications (Table 2).

**Fig. 4.**
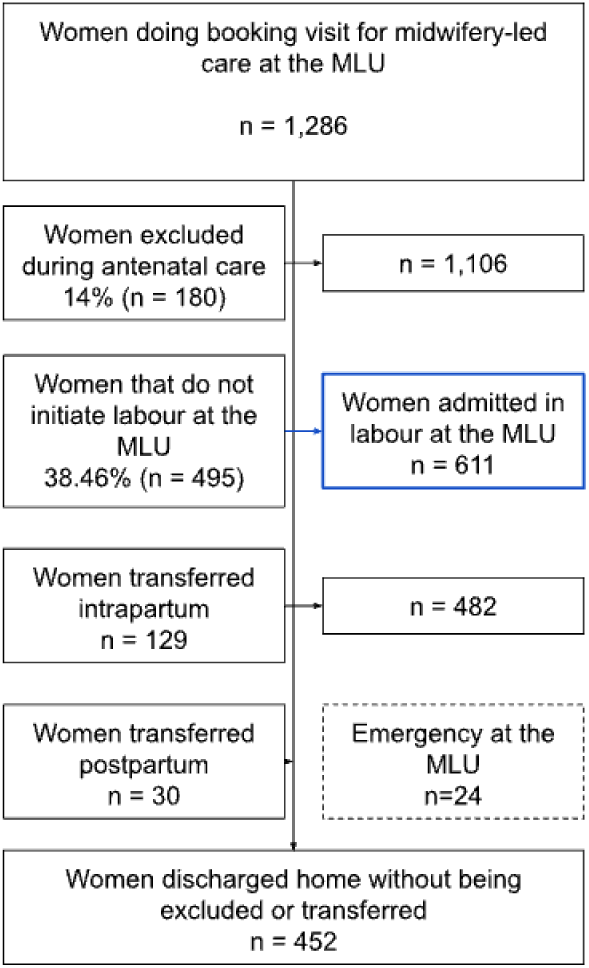
Pathway of care for women in the MLU

**Table 2.**
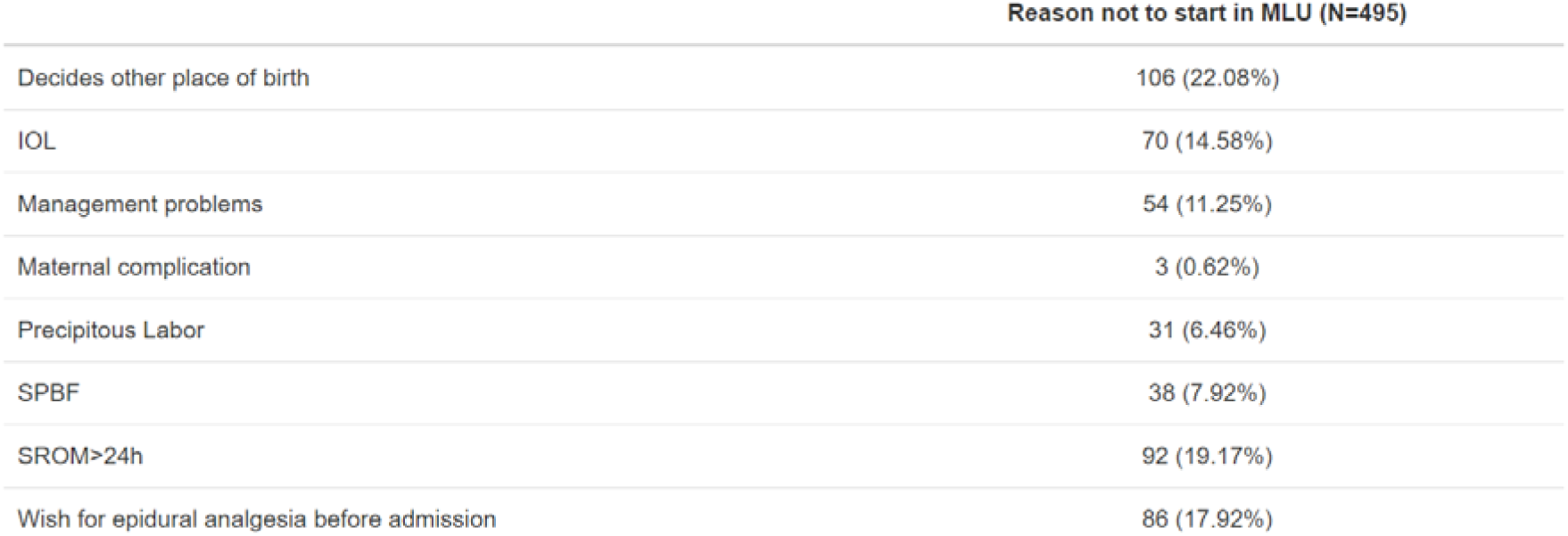
Reasons for not being admitted to the MLU at the onset of labour.

| Reason not to start in MLU (N=495) |  |
| --- | --- |
| Decides other place of birth | 106 (22.08%) |
| IOL | 70 (14.58%) |
| Management problems | 54 (11.25%) |
| Maternal complication | 3 (0.62%) |
| Precipitous Labor | 31 (6.46%) |
| SPBF | 38 (7.92%) |
| SROM>24h | 92 (19.17%) |
| Wish for epidural analgesia before admission | 86 (17.92%) |

As for women who did initiate labour at the MLU, only 4.1% required caesarean sections (Figure 5), well below the percentages of HM and other level-I hospitals (Figure 1), while 92.3% had spontaneous births (Figure 5). Nevertheless, the MLU experienced similar trends in terms of types of birth to those observed for HM (Figure 1): caesarean sections generally decreased from 2018 to 2021 and rose again in 2022, although a modest increase was observed in 2020; and spontaneous births showed the opposite pattern. Moreover, from 2019 onwards, an exponentially growing proportion of such spontaneous births involved water immersion (Figure 5).

**Fig. 5.**
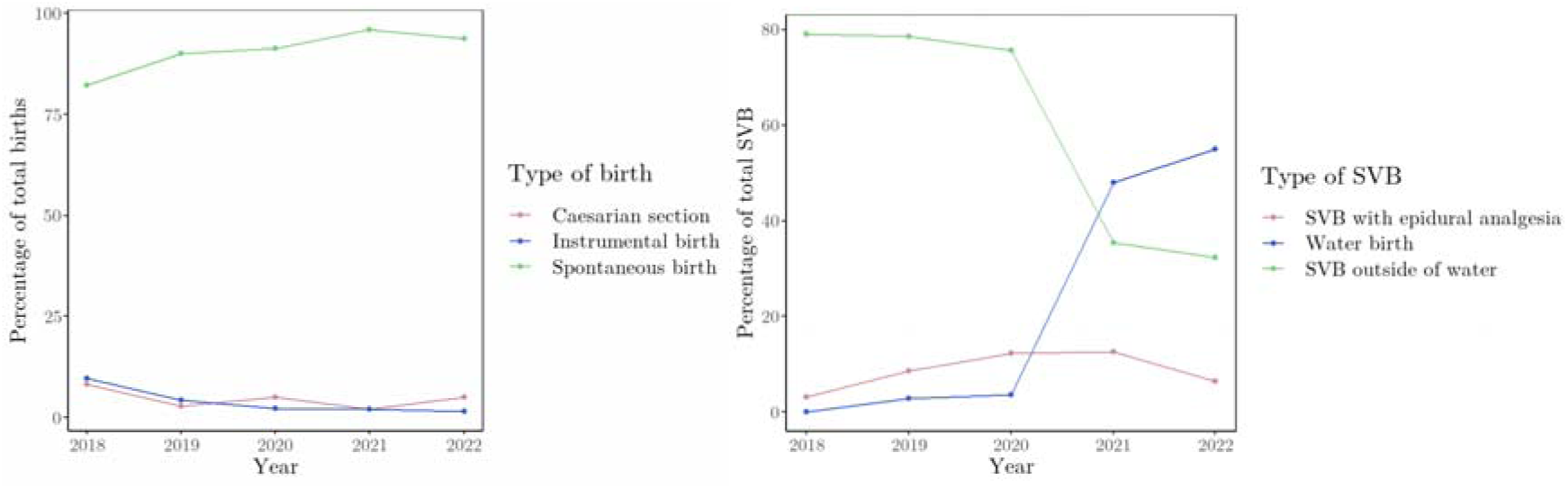
Type of birth for women in the MLU and characteristics of spontaneous vaginal births.

Throughout the study period, only 24 emergencies were registered in the MLU (Figure 4). Of these, 10 involved neonatal resuscitation (NNR), 6 were due to shoulder dystocia, 6 due to postpartum haemorrhage (PPH), and 2 involved both NNR and PPH. Focusing on the newborns implied in these 24 emergencies, 7 of them scored an Apgar <7, and 6 had to be transferred to neonatal intensive care units (NICU).

Beyond these emergencies, 129 of the 611 women who initiated active labour in the MLU required intrapartum transfers, and an additional 30 needed to be transferred postpartum (Figure 4). Notably, the share of total transfers remained fairly stable over the 5 years (Supplementary Table 5). Of all these women, only 10.3% of multiparous women had to be transferred, whereas in the case of nulliparous women, 39% required transfer. (Supplementary Table 6).

The most prevalent reasons for intrapartum transfer were the desire for epidural analgesia (51.9%) and delayed first stages of labour (21.7%). Other reasons were meconium-stained liquor (9.3%), delayed second stages of labour (7.0%), abnormal foetal heart rate (7.0%), and maternal complications (0.8%) (Supplementary table 7).

## 4. DISCUSSION

This study aimed to evaluate the implementation and impact of the first alongside MLU in Spain, a country with a highly medicalised approach to childbirth, over its first five years of operation. The introduction and subsequent closure of HM’s MLU provide a unique case study within the Catalan healthcare system. In that regard, the findings highlight how the existence of a MLU altered the pattern of childbirth interventions at HM. Specifically, the introduction of the MLU at HM led to a marked increase in spontaneous vaginal births and a corresponding decrease in caesarean sections, aligning with the WHO’s recommended caesarean sections rates of 10-15% (WHO, 2015). This trend not only applied to women admitted in labour at the MLU, where spontaneous vaginal births constituted the majority of deliveries. The implementation of the MLU also led to an improvement in the share of these births over time in HM’s OU, which may be attributed to women transferred from the MLU being less likely to receive interventions (Palau-Costafreda et al., 2023). Meanwhile, other level-I hospitals in Catalonia experienced a slight increase in caesarean sections, in line with the alarming projections by Bertran and colleagues (2021), who estimated global caesarean section rates to reach 28.5% by 2030 and 36.5% in Europe. However, with the closure of the MLU, the trends in childbirth interventions reversed, with spontaneous vaginal births dropping to percentages similar to those at other level-I hospitals. Therefore, these findings highlight the beneficial impact of MLUs on promoting physiological childbirth and reducing intervention rates (Sandall et al., 2018; Scarf et al., 2018; Yu et al., 2020), while emphasising the critical need for sustained support and investment in such units to maintain positive outcomes in time.

Beyond childbirth intervention trends, women’s interest in the MLU nearly tripled from 2018 to 2022, peaking at 2021. The COVID-19 pandemic likely contributed to this peak, as women contingently sought personalised, less crowded childbirth environments (Flaherty et al., 2022; Hadjigeorgiou et al., 2022; Mortazavi & Ghardashi, 2021; Preis et al., 2021). However, this tendency could also be attributed to increasing confidence in midwifery-led care and a growing preference for less medicalised birth settings (Newburn, 2003). Qualitative accounts of women who gave birth at MLUs suggest that these units embody a model of care that promotes positive childbirth experiences (Palau-Costafreda et al., forthcoming). Research also shows that women’s decisions about where to give birth are significantly influenced by personal research and recommendations from friends and online communities (Yuill et al., 2020). Therefore, the positive childbirth experiences of women attended at HM’s MLU likely attracted more women. This idea is further supported by the steady increase in the number of women from outside HM’s coverage area interested in the MLU throughout the study period. It should still be noted that this interest did not translate into a greater number of births attended at the MLU. A substantial proportion of these women ultimately chose not to give birth at the MLU due to geographical accessibility constraints. As such, establishing additional MLUs could enable more women to opt for midwifery-led care without having to compromise their preference due to distance (Hermus et al., 2015). Lastly, the fact that women’s interest in the MLU stalled in 2022 was possibly related to saturation in awareness and acceptance of the MLU, the opening of more MLUs in the area (Alcaraz et al., 2024), or the organisational challenges that led to the unit’s closure at the end of 2022, as women potentially sought more stable and long-term options for their childbirth care (Pelak, Dahlen & Keedle, 2023). Indeed, with the closure of the MLU, fewer women chose HM for childbirth, indicating a clear preference for the supportive environment provided by the MLU.

The increase in women with higher initial pregnancy risk assessments interested in the MLU over the years reflects ongoing updates to the MLU’s inclusion criteria, aligning with national (Generalitat de Catalunya, 2018) and international guidelines (NICE, 2014). This evolution highlights the unit’s commitment to adhering to evidence-based practices and empowering women to make informed choices, which are key strategies for promoting normality in childbirth (Kennedy et al., 2010; Renfrew et al., 2014) and achieving lower childbirth intervention rates (He et al., 2023). An insightful finding is the increasing prevalence of water births in 2021, when the MLU introduced new protocols and staff training to attend water births. That year, water births accounted for 48.0% of spontaneous vaginal births, increasing to 54.9% in 2022, indicating a strong preference for water births among women at the MLU once the option became available. Again, the ability to adapt its practices, both in the MLU and the OU, reflects its dedication to providing high-quality, patient-centred care, aligning with best practices in maternity care globally (Renfrew et al., 2014).

Results also demonstrate the safety and effectiveness of HM’s MLU. The incidence of emergencies in the MLU was low (2.9%), consistent with findings from high-income countries (Scarf et al., 2018). Neonatal admission to the NICU during the study period was 1.3%, a rate comparable to studies from countries where MLUs are well implemented, including Sweden (Gottvall et al., 2005), England (Birthplace in England Collaborative Group, 2011) or Australia (Homer et al., 2014). The average transfer rate from the MLU to the OU during labour or immediately after birth was 26%, with nulliparous women being transferred more frequently than multiparous women. These transfer rates are also comparable to those reported in England (Birthplace in England Collaborative Group, 2011). Moreover, most transfers were non-urgent, the most common reason being the desire for epidural analgesia. Therefore, the stable transfer rates and effective management of emergencies suggest that HM’s MLU was successfully integrated into the healthcare system, providing safe and supportive care for a significant number of women. In any case, these findings still point out the importance of immediate and effective emergency protocols, continuous training for staff, and interprofessional collaboration.

### Implications for practice

The findings from HM’s MLU highlight several implications for maternity care in Spain. The positive outcomes observed suggest that further integration of MLUs within the Spanish National Health System could contribute to reducing unnecessary medical interventions and promoting physiological childbirth processes. This is crucial to improve the overall quality of maternal care and align with best practices recommended by the WHO and other health authorities. However, for the midwifery-led care model to be sustainable and effective, support is needed in various key areas. Proper resource allocation, including necessary equipment and facilities, is essential for the efficient and effective operation of MLUs. Additionally, ongoing education and professional development for midwives are critical to continuously implementing evidence-based practices and maintaining high standards of care (Pelak, Dahlen & Keedle, 2023). Finally, increasing public awareness about the benefits of MLUs and ensuring accessibility for all women, regardless of their geographical location, is essential to maximise their impact and avoid inequities in women’s access to midwifery-led care (Fontenot et al., 2024; Scarf et al., 2018).

The potential economic benefits of integrating MLUs into the healthcare system are also considerable. According to a 2019 report by the Catalan Department of Health (Generalitat de Catalunya, 2019), spontaneous vaginal births incur fewer costs compared to instrumental vaginal births and caesarean sections. Women who undergo spontaneous vaginal births generally require fewer visits to primary care services, emergency hospital visits, and outpatient consultations during the first three months postpartum, which translates into cost savings for the healthcare system. Additionally, the report highlights that women who experience instrumental vaginal births or caesarean sections have higher rates of medication dispensation, including analgesics, antibiotics, and antidepressants, particularly during the first three months postpartum. These costs persist throughout the first year for women who undergo caesarean sections, further increasing the financial burden on the healthcare system. However, the current financing model of the Catalan healthcare system, which funds a pre-set fee based on the hospital’s level of complexity regardless of the type of births attended, hampers comprehensive studies to capture the cost differences associated with different birthing methods. Therefore, integrating cost-effectiveness analyses into policy decision-making processes could lead to more nuanced and efficient resource allocation, potentially incentivising the adoption of MLUs.

All these factors combined make a compelling case for the spread and continuous support of MLUs within the Catalan and Spanish healthcare system.

### Limitations and strengths

This study has several strengths and limitations. One significant strength is the combination of individual-level data from HM’s MLU and population-based data drawn on the MBDS of the Catalan Health Service. This reliable dataset allows for detailed comparisons and trend analysis across various hospital levels over a significant period (2018-2023), providing valuable insights into the impacts of the MLU on childbirth outcomes. The comparison between HM and other level I, II, and III hospitals enhances the study’s representativeness. Further, as the first study to overview childbirth interventions in Catalonia since MLUs were implemented, it offers valuable baseline data and contributes significantly to the knowledge of midwifery-led care in traditionally obstetrician-led systems. However, the findings may have limited generalizability beyond Catalonia due to the decentralised Spanish healthcare structure, cultural factors, and contingent factors specific to the implementation of the MLU within HM. That is, the study period includes the COVID-19 pandemic, which influenced MLU operations and general childbirth trends. The pandemic’s impact on healthcare services, patient preferences, and hospital operations could confound the study’s findings during that period of time.

Future research should include larger prospective studies across multiple MLUs to validate these findings and provide more insights into the long-term impact of midwifery-led care in Spain. Understanding the high demand for MLU services and comparing experiences with traditional obstetric care could further improve maternity services.

## 5. CONCLUSION

Lessons from the implementation of the first MLU in Spain point out the significant impact that MLUs can have on reducing medical interventions during childbirth. This particular case demonstrates that such units can lead to lower caesarean section rates and higher spontaneous vaginal birth rates, contributing to more positive maternal and neonatal outcomes. However, the closure of the MLU and the subsequent regression in HM’s birth outcomes highlight the importance of sustained support and investment in these units. In addition to childbirth intervention trends, the incidence of emergencies within the MLU was low and comparable to rates observed in countries where MLUs are well established. This indicates that the implementation of the MLU was successful and able to provide safe and effective care. To this should be added that HM’s MLU demonstrated a strong commitment to continuous improvement and adherence to evidence-based best practices. This commitment ensured that the unit remained updated with the latest guidelines and recommendations, thereby enhancing the quality of care provided to women and their newborns. Beyond birth outcomes, there is clear interest from women in this model of care, as shown by an increasing demand for MLU services over the 5-year period. Policymakers and healthcare providers should therefore consider these findings as they work towards creating a more efficient, effective, and compassionate maternity care system in Spain.

## Data Availability

All data produced in the present study are available upon reasonable request to the authors

## Footnotes

1 Considering that fieldwork started in December 2017, only 4 women were registered in the dataset that year. Therefore, in this section they will be merged with women that were attended in 2018.

