## Supplementary material for "Impact of Midwifery-led units in Spain: lessons from the first 5 years"

| Births by level of care in Catalonia | | | | | | | | | | | | | | | | | | |
|---|---|---|---|---|---|---|---|---|---|---|---|---|---|---|---|---|---|---|
| Year | 2018 | | | 2019 | | | 2020 | | | 2021 | | | 2022 | | | 2023 | | |
| Level of care | I | II | III | I | II | III | I | II | III | I | II | III | I | II | III | I | II | III |
| | N = 12121 | N = 13421 | N = 17272 | N = 11705 | N = 13511 | N = 17522 | N = 10154 | N = 12387 | N = 17835 | N = 9681 | N = 12167 | N = 17468 | N = 10153 | N = 12125 | N = 16958 | N = 9610 | N = 11753 | N = 17269 |
| Caesarian section | 2348 (19.37%) | 2672 (19.91%) | 4301 (24.90%) | 2410 (20.59%) | 2785 (20.61%) | 4452 (25.41%) | 2020 (19.89%) | 2623 (21.18%) | 4589 (25.73%) | 2020 (20.87%) | 2673 (21.97%) | 4532 (25.94%) | 2230 (21.96%) | 2699 (22.26%) | 4355 (25.68%) | 2063 (21.47%) | 2658 (22.62%) | 4707 (27.26%) |
| Spontaneous birth | 8493 (70.07%) | 9083 (67.68%) | 11107 (64.31%) | 8122 (69.39%) | 9150 (67.72%) | 11104 (63.37%) | 7114 (70.06%) | 8411 (67.90%) | 11391 (63.87%) | 6686 (69.06%) | 8153 (67.01%) | 11310 (64.75%) | 6937 (68.32%) | 8072 (66.57%) | 10994 (64.83%) | 6592 (68.60%) | 7836 (66.67%) | 11060 (64.05%) |
| Instrumental birth | 1280 (10.56%) | 1666 (12.41%) | 1864 (10.79%) | 1173 (10.02%) | 1576 (11.66%) | 1966 (11.22%) | 1020 (10.05%) | 1353 (10.92%) | 1855 (10.40%) | 975 (10.07%) | 1341 (11.02%) | 1626 (9.31%) | 986 (9.71%) | 1354 (11.17%) | 1609 (9.49%) | 955 (9.94%) | 1259 (10.71%) | 1502 (8.70%) |

| | Total births HM | | | | | | |
|---|---|---|---|---|---|---|---|
| | Total | 2018 | 2019 | 2020 | 2021 | 2022 | 2023 |
| | N = 3775 | N = 625 | N = 612 | N = 698 | N = 728 | N = 702 | N = 410 |
| Caesarian section | 666 (17.64%) | 147 (23.52%) | 100 (16.34%) | 106 (15.19%) | 98 (13.46%) | 121 (17.24%) | 94 (22.93%) |
| Spontaneous birth | 2766 (73.27%) | 401 (64.16%) | 443 (72.39%) | 536 (76.79%) | 573 (78.71%) | 530 (75.50%) | 283 (69.02%) |
| Instrumental birth | 343 (9.09%) | 77 (12.32%) | 69 (11.27%) | 56 (8.02%) | 57 (7.83%) | 51 (7.26%) | 33 (8.05%) |

| | Total | 2017-2018 | 2019 | 2020 | 2021 | 2022 |
|---|---|---|---|---|---|---|
| | *N=611* | *N=62* | *N=70* | *N=139* | *N=198* | *N=142* |
| Type of birth: | | | | | | |
| Caesarian section | 25 (4.09%) | 5 (8.06%) | 2 (2.86%) | 7 (5.04%) | 4 (2.02%) | 7 (4.93%) |
| Spontaneous birth | 564 (92.3%) | 51 (82.3%) | 63 (90.0%) | 127 (91.4%) | 190 (96.0%) | 133 (93.7%) |
| Instrumental birth | 18 (2.95%) | 6 (9.68%) | 3 (4.29%) | 3 (2.16%) | 4 (2.02%) | 2 (1.41%) |

| | Total | 2017-2018 | 2019 | 2020 | 2021 | 2022 |
|---|---|---|---|---|---|---|
| | *N=564* | *N=51* | *N=63* | *N=127* | *N=190* | *N=133* |
| Type of spontaneous vaginal birth (SVB): | | | | | | |
| SVB with epidural analgesia | 59 (10.5%) | 2 (3.92%) | 6 (9.52%) | 17 (13.4%) | 25 (13.2%) | 9 (6.77%) |
| SVB outside of water | 325 (57.6%) | 49 (96.1%) | 55 (87.3%) | 105 (82.7%) | 70 (36.8%) | 46 (34.6%) |
| Water birth | 180 (31.9%) | 0 (0.00%) | 2 (3.17%) | 5 (3.94%) | 95 (50.0%) | 78 (58.6%) |

| | Total | 2017-2018 | 2019 | 2020 | 2021 | 2022 |
|---|---|---|---|---|---|---|
| | *N=592* | *N=59* | *N=64* | *N=135* | *N=193* | *N=141* |
| Transfer: | | | | | | |
| No | 433 (73.1%) | 43 (72.9%) | 47 (73.4%) | 99 (73.3%) | 142 (73.6%) | 102 (72.3%) |
| Yes | 159 (26.9%) | 16 (27.1%) | 17 (26.6%) | 36 (26.7%) | 51 (26.4%) | 39 (27.7%) |

| | Total | Nulliparous | Multiparous |
|---|---|---|---|
| | *N=587* | *N=344* | *N=243* |
| Transfer: | | | |
| No | 428 (72.9%) | 210 (61.0%) | 218 (89.7%) |
| Yes | 159 (27.1%) | 134 (39.0%) | 25 (10.3%) |

| | Total | 2017-2018 | 2019 | 2020 | 2021 | 2022 |
|---|---|---|---|---|---|---|
| | *N=129* | *N=15* | *N=15* | *N=32* | *N=42* | *N=25* |
| Reason for intrapartum transfer: | | | | | | |
| Wish for epidural analgesia | 67 (51.9%) | 6 (40.0%) | 5 (33.3%) | 21 (65.6%) | 23 (54.8%) | 12 (48.0%) |
| Delayed 1st stage | 28 (21.7%) | 6 (40.0%) | 4 (26.7%) | 3 (9.38%) | 10 (23.8%) | 5 (20.0%) |
| Meconium stained liquor | 12 (9.30%) | 1 (6.67%) | 1 (6.67%) | 3 (9.38%) | 6 (14.3%) | 1 (4.00%) |
| Abnormal fetal heart rate | 9 (6.98%) | 0 (0.00%) | 0 (0.00%) | 3 (9.38%) | 2 (4.76%) | 4 (16.0%) |
| Delayed 2nd stage | 9 (6.98%) | 1 (6.67%) | 3 (20.0%) | 1 (3.12%) | 1 (2.38%) | 3 (12.0%) |
| Maternal complication | 1 (0.78%) | 0 (0.00%) | 1 (6.67%) | 0 (0.00%) | 0 (0.00%) | 0 (0.00%) |
